# Patient-Reported Reasons for Antihypertensive Medication Change: A Quantitative Study Using Social Media

**DOI:** 10.1101/2023.08.01.23293490

**Authors:** Cristina Micale, Su Golder, Karen O’Connor, Davy Weissenbacher, Robert Gross, Sean Hennessy, Graciela Gonzalez-Hernandez

## Abstract

**Introduction:** Hypertension is the leading cause of heart disease in the world, and discontinuation or nonadherence of antihypertensive medication constitutes a significant global health concern. Patients with hypertension have high rates of medication nonadherence [13–15]. Studies of reasons for nonadherence using traditional surveys are limited, can be expensive, and suffer from response, white-coat, and recall biases. Mining relevant posts by patients on social media is inexpensive and less impacted by the pressures and biases of formal surveys, which may provide direct insights into factors that lead to non-compliance with antihypertensive medication.

**Methods:** This study examined medication ratings posted to WebMD, an online health forum that allows patients to post medication reviews. We used a previously developed natural language processing classifier to extract indications and reasons for changes in angiotensin receptor II blocker (ARBs) and angiotensin-converting enzyme inhibitor (ACEIs) treatments. After extraction, ratings were manually annotated and compared to data from the FAERS public database.

**Results:** From a collection of 343,459 WebMD reviews, we automatically extracted 1,867 posts mentioning changes in ACEIs or ARBs, and manually reviewed the 300 most recent posts regarding ACEI treatments and the 300 most recent posts regarding ARB treatments. After excluding posts that only mentioned a dose change or were a false positive mention, 142 posts in the ARBs dataset and 187 posts in the ACEIs dataset remained. The majority of posts (97% ARBs, 91% ACEIs) indicated experiencing an adverse event as the reason for medication change. The most common adverse events reported mapped to the Medical Dictionary for Regulatory Activities were “musculoskeletal and connective tissue disorders” like muscle and joint pain for ARBs, and “respiratory, thoracic, and mediastinal disorders” like cough and shortness of breath for ACEIs. These categories also had the largest differences in percentage points, appearing more frequently on WebMD data than FDA data (p=0.000).

**Conclusion:** Musculoskeletal and respiratory symptoms were the most commonly reported adverse effects in social media postings associated with drug discontinuation. Managing such symptoms is a potential target of interventions seeking to improve medication persistence.

**Key Points:**

1. The major reason for the discontinuation of ARBs and ACEIs expressed through WebMD was the experience of adverse events.
2. Musculoskeletal and respiratory symptoms were the most commonly reported adverse effects in social media postings associated with drug discontinuation.
3. This study shows the promise of WebMD mining as an effective tool in medication nonadherence and adverse event research.

## 1 Introduction

### 1.1 Medication Nonadherence and Discontinuation Background

Medication nonadherence is a global issue in healthcare that is difficult to study and control. Nonadherence refers to when a patient does not follow a healthcare provider’s instructions for taking a certain medication [1–3, 5]. This includes taking too little or too much of the medication, stopping the medication without the consultation of a healthcare provider (referred to as non-persistence), or not starting the treatment at all. Studies have concluded that medication nonadherence costs the United States healthcare system more than $200 billion yearly [2]. Additionally, compared to those who are adherent, patients over the age of 50 who are nonadherent have a 17% higher risk of hospitalization for any cause [3]. Although the effects of nonadherence are evident, the causes of nonadherence are not well studied.

Nonadherence and discontinuation of medication is a particular challenge for the treatment of chronic diseases [4–7]. There are higher degrees of adherence for a treatment that cures an illness (77% adherence) when compared to a treatment that prevents an illness (63% adherence), and long treatments prompt a reduction in adherence to about 50% [10]. A meta-analysis studying persistence in diabetes and hypertension treatments found that (1) only approximately 63% of chronic condition patients continue their treatment for a year, and (2) patients follow their treatment plan only 72% of the time it is prescribed for [4]. Some patients may consciously fail to adhere to their treatment because of adverse drug events, a belief that the medication is not working, a belief that they do not need the medication any more, or high drug costs [1, 10, 11]. Moreover, lack of understanding of their treatment may contribute to both intentional and unintentional nonadherence. Because the success of chronic disease treatments depends on consistency [7–9], it is important to determine why patients discontinue their medication, and even more so when they discontinue medication without the supervision of their healthcare provider.

### 1.2 Antihypertensive Medication Nonadherence

Hypertension is the leading cause of heart disease in the world [12–15], and 43-65.5% of patients who are non adherent to their chronic medication regimens are hypertensive patients [13–15]. Nonadherence to antihypertensive medications is especially dangerous for patients with various comorbidities or for those with uncontrolled blood pressure (systolic over 140 mmHg and diastolic over 90 mmHg) [12]. Reducing blood pressure by 10 mmHg significantly reduces the risk of stroke, CVD, CHD, heart failure, diabetes, and kidney disease; therefore, it is important to understand why hypertensive patients are continuously not adhering to their medication regimens [12, 17]. A previous study analyzing initial medication nonadherence (IMN) found that 24% of patients in the United States did not even pick up their antihypertensive prescriptions due to five main reasons in order of prevalence: (1) cost, (2) concern for side effects, (3) other concerns for taking the medication, (4) health insurance, and (5) thinking that the medication was not necessary [16].

It has been shown that discontinuation of antihypertensive medication can be done to conduct certain diagnostic examinations [8] or washout periods in clinical trials [9]. Certain patients can remain normotensive after discontinuation of their medication, and the risk of acute cardiovascular and cerebrovascular adverse events can remain low in a short-term period. However, these discontinuations must be well-controlled and blood pressure must be carefully tracked throughout the discontinuation period. Furthermore, withdrawal of antihypertensive treatment may only be successful for mild to moderate hypertension or for patients who will be complying to drastic lifestyle changes for the long-term [18]. These cases are not the most common, yet a high percentage of hypertensive patients discontinue their medication [13–15]; therefore, patient perspectives and possible misinformation about antihypertensive medication discontinuation are important to study.

### 1.3 Social Media and Studying Medication Nonadherence

Traditional epidemiological studies of medication nonadherence have focused on measuring the rates and outcomes of nonadherence, not the reasons behind these outcomes. Moreover, these studies have relied on the data from direct tests, clinical records, pharmacy claims, pill counts, and patient surveys [2, 14, 19, 24]. These sources only provide a very small fraction of the nonadherence occurring and may be influenced by response bias, white coat bias, and/or recall bias [2, 19, 24]. Self-reporting scales have a variety of disadvantages: some are too time-consuming, others have limited generalizability, and many rely on patients having high literacy and proficient recall of their adherence [24]. One 2007 alternative-method study determined that angiotensin-converting enzyme (ACE) inhibitor adherence could be verified by a serum ACE activity test, which revealed that 58% of 73 chronic heart failure patients were non-adherent to their ACE inhibitor treatment [20]. Although this serum test was inexpensive, it required participants to be patients who regularly attended a clinic or hospital to be able to track non-adherence [20]. Because ACE inhibitors are the most commonly prescribed medications to treat cardiovascular diseases like hypertension [21], this is not a feasible or efficient way of studying general ACE inhibitor nonadherence. Angiotensin receptor II blockers are commonly prescribed as alternatives to ACE inhibitors [22], and no discrete exam has been developed to verify discontinuation or nonadherence. Nonetheless, any methods involving direct contact with healthcare personnel introduces white coat adherence, a phenomenon where patient adherence to treatments increases by as much as 20% around clinic visits or tests [24]. Overall, it is evident that medication nonadherence research has been restricted to focusing on outcomes and been heavily influenced by bias because of traditional methodology.

Social media platforms, such as Twitter, have been a new pathway that can reflect a complementary picture of reasons for nonadherence, reducing recall bias (inaccuracies in patient’s retrieval of information from the past), white coat bias (inaccuracies in patient responses due to anxiety about doctor visits) and response bias (general inaccuracies or falsity in participant responses to structured interviews or surveys). Many studies have started to extract data from Twitter to analyze drug-switching, patient perspectives on medications, and mentions of nonadherence [2, 23]. These methods use first-hand, easily accessible data to provide understanding of patient sentiments, perspectives, and experiences. Recent studies have also used these automated methods to identify mentions of nonadherence on social media platforms and online drug review sites such as WebMD [23, 25]. Patients can write reviews for specific medications on this platform, and many contain active mentions of nonadherence. Golder et al. studied nonadherence in statin medications and the reasons behind it [25]. The goal of this study is to conduct a similar investigation to understand patients’ reasons behind nonadherence and discontinuation of antihypertensive medication. This data may provide useful information to help decrease antihypertensive medication nonadherence rates, which will in turn help improve the outcomes of hypertension treatment in the United States.

## 2 Methods

Two classes of antihypertensive medications, ACE inhibitors (ACEIs) and angiotensin II receptor blockers (ARBs), were chosen because they are two of the most prescribed medications in the United States for hypertension [21, 22]. Using all the available generic and brand names for ACEIs and ARBs (see Supplementary Materials Table 1), we retrieved 6,085 reviews (4,545 ARBs, 1540 ACEIs) from our WebMD collection of reviews posted from January 1, 2008 - September 9, 2021. Each post contains the comment, date, author age and gender, time on the drug, author role, condition, overall rating, effectiveness, ease of use, and satisfaction rating. An existing trained classifier that identifies WebMD posts indicating a change in medication treatment that has been validated to operate with 0.871 precision was used in this study [26]. The classifier identified 1,867 reviews as containing an indication of a treatment change.

Of the 1867 posts, 1418 were for ARBs and 449 were for ACEIs. The posts were ordered by recency, and the most recent 300 for each medication category were manually annotated to identify nonpersistence and characterize the reasons for it whenever the user listed it. For each review, the annotators coded for the type of medication change (“discontinue”, “switch”, “dosage change”, or “none”), whether the author of the review appears to have had a healthcare consultation for the medication change (“yes,” “no,” “unsure”), whether the change was patient or healthcare directed, the reason for the change (“adverse events,” “cost/insurance,” “not working,” “not necessary,” or “other”), plus, if applicable, adverse events mentioned, the number of days before the emergence of adverse events, whether the author mentions rechallenge of the medication, and whether the adverse events returned after the rechallenge. Adverse events were extracted manually, and were coded to the appropriate System Organ Class (SOC) level Medical Dictionary for Regulatory Activities (MedDRA) terms upon analysis of the data. Two annotators, both experienced in social media data annotation for at least two years, developed the aforementioned annotation guidelines through a series of revisions with input from other social media NLP research experts. A random sample of 10% of the posts were double annotated, and the inter-annotator agreement was high.

Figure 1 shows an example of the annotation data collected for a post on losartan potassium that was characterized as non-persistence because the comment indicates a discontinuation of medication that was patient directed and a direct mention of not having a health care consultation for the mentioned change.

**Figure 1.**
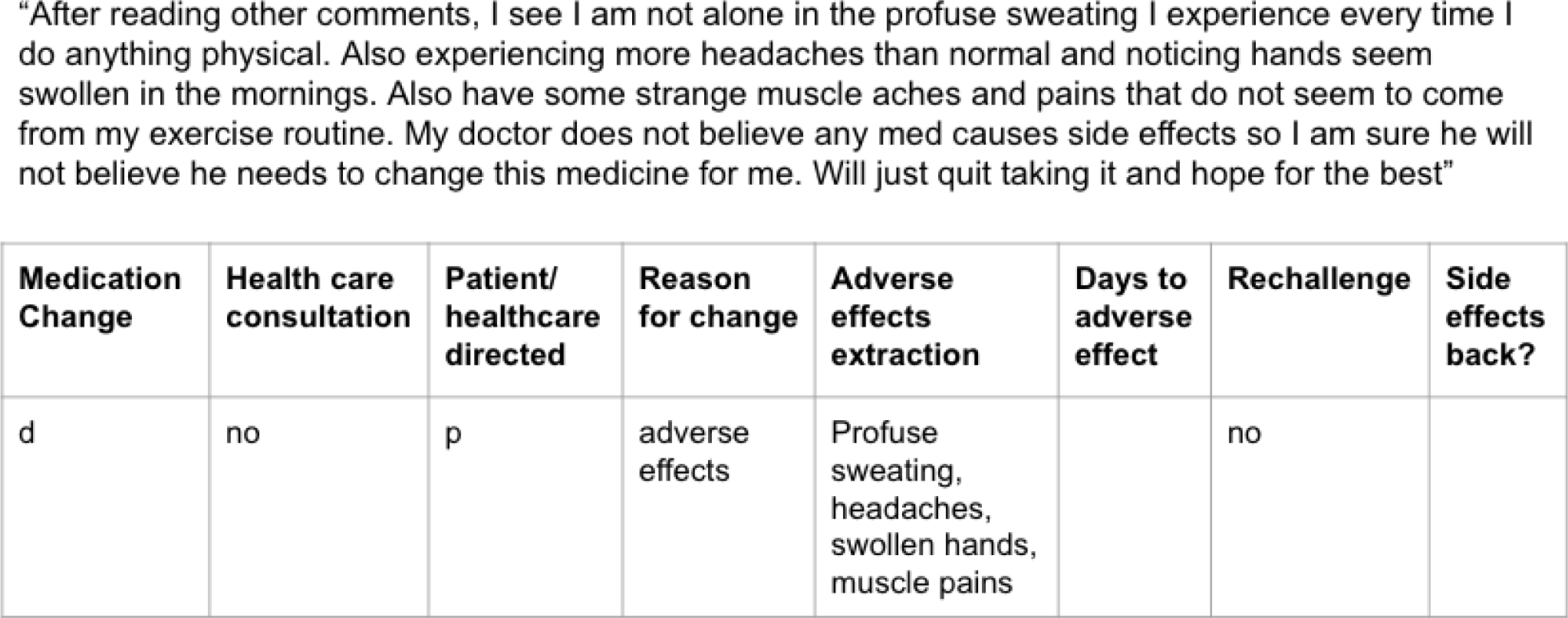
Sample annotation for losartan potassium indicating nonadherence.

FDA reports of adverse events regarding each medication class were also collected through the FDA Adverse Events Reporting System (FAERS) public database. From 1968 to 2022, a total of 759,742 reports of ARBs and 512,196 reports of ACEIs from the FAERS database were analyzed, and the SOCs were compared to the WebMD results through the percentage point difference statistical test. Similar data was collected and superficially analyzed from the Medicines and Healthcare products Regulatory Agency (MHRA) database. MHRA data offered similar results to FDA data, and because a minimal percentage of WebMD posts come from users in the United Kingdom, MHRA data was only briefly included in the results figures.

## 3 Results

### 3.1 Type of Medication Change and Patient Demographics

Of the original corpus, 600 total posts were annotated (inter-annotator agreement was 0.9 as most of the data was not subjective) and only those indicating a discontinuation or switch to another medication were included in the analysis of nonadherence. In the ARBs set, 37% (110/300) of posts were excluded because they did not indicate a medication change and 16% (48/300) were excluded because they only indicated a dosage change. In the ACEIs set, 29% (86/300) of posts were excluded because they did not indicate a change and 9% (27/300) only indicated a dosage change. Of the remaining posts, 16% (49/300) of ARBs posts and 22% (65/300) of ACEIs posts indicated a medication switch and 31% (93/300) of ARBs and 41% (122/300) of ACEIs posts indicated a discontinuation of medication.

41% (58/142) of the patients in the ARBs dataset and 56% (104/187) in the ACEIs dataset self-reported female, 21% (30/142) of ARBs patients and 32% (60/187) of ACEIs patients self-reported male, and 38% (54/142) of ARBs posts and 12% (23/187) of ACEIs posts did not declare their biological sex through their WebMD post. The most common age range categories were 45-54, 55-64, and 65-74 encompassing 85% of ARBs posts and 82% of ACEIs posts. Additionally, most patients (51% ARBs and 52% ACEIs) had been taking their antihypertensive medication for less than six months.

### 3.2 Reasons for Discontinuation/Switching

The overwhelming reason for discontinuation or a medication switch was the experience of adverse events (138/142, 97% ARBs; 170/187, 91% ACEIs), followed by the medication not working (13/142, 9% ARBs; 6/187, 3% ACEIs) and cost/insurance reasons (7/142, 5% ARBs; 7/187, 4% ACEIs). Most mentions of the medication not working were associated with a mention of an adverse event.

After adverse events were extracted from the comments, they were coded to MedDRA SOC organ classes. The most mentioned MedDRA categories in the ARBs dataset were ‘Musculoskeletal and connective tissue disorders’ (71/396, 18%), ‘Nervous system disorders’ (68/396, 17%), and ‘General disorders and administration site conditions’ (56/396, 14%). The most mentioned categories in the ACEIs posts were ‘Respiratory, thoracic and mediastinal disorders’ (114/435, 26%), ‘Nervous system disorders’ (69/435, 16%), and ‘Gastrointestinal disorders’ (61/435, 14%).

WebMD adverse event reports were then compared to the FDA reports extracted from the FAERS public database (Figures 2 and 3). The ‘General disorders and administration site conditions’, ‘Skin and subcutaneous tissue disorders,’ ‘Investigations,’ ‘Vascular disorders,’ ‘Metabolism and nutrition disorders,’ ‘Renal and urinary disorders,’ ‘Injury, poisoning and procedural complications,’ and ‘Cardiac disorders’ MedDRA SOC categories were mentioned more frequently in FDA reports than in WebMD posts for both medication classes. However, a larger proportion of ARBs posts on WebMD mentioned adverse events related to ‘Musculoskeletal and connective tissue disorders’ when compared to ARBs FDA report proportions. Similarly, the WebMD reports for ACEIs showed a greater proportion of mentions of ‘Respiratory, thoracic and mediastinal disorders’ than FDA reports.

**Figure 2.**
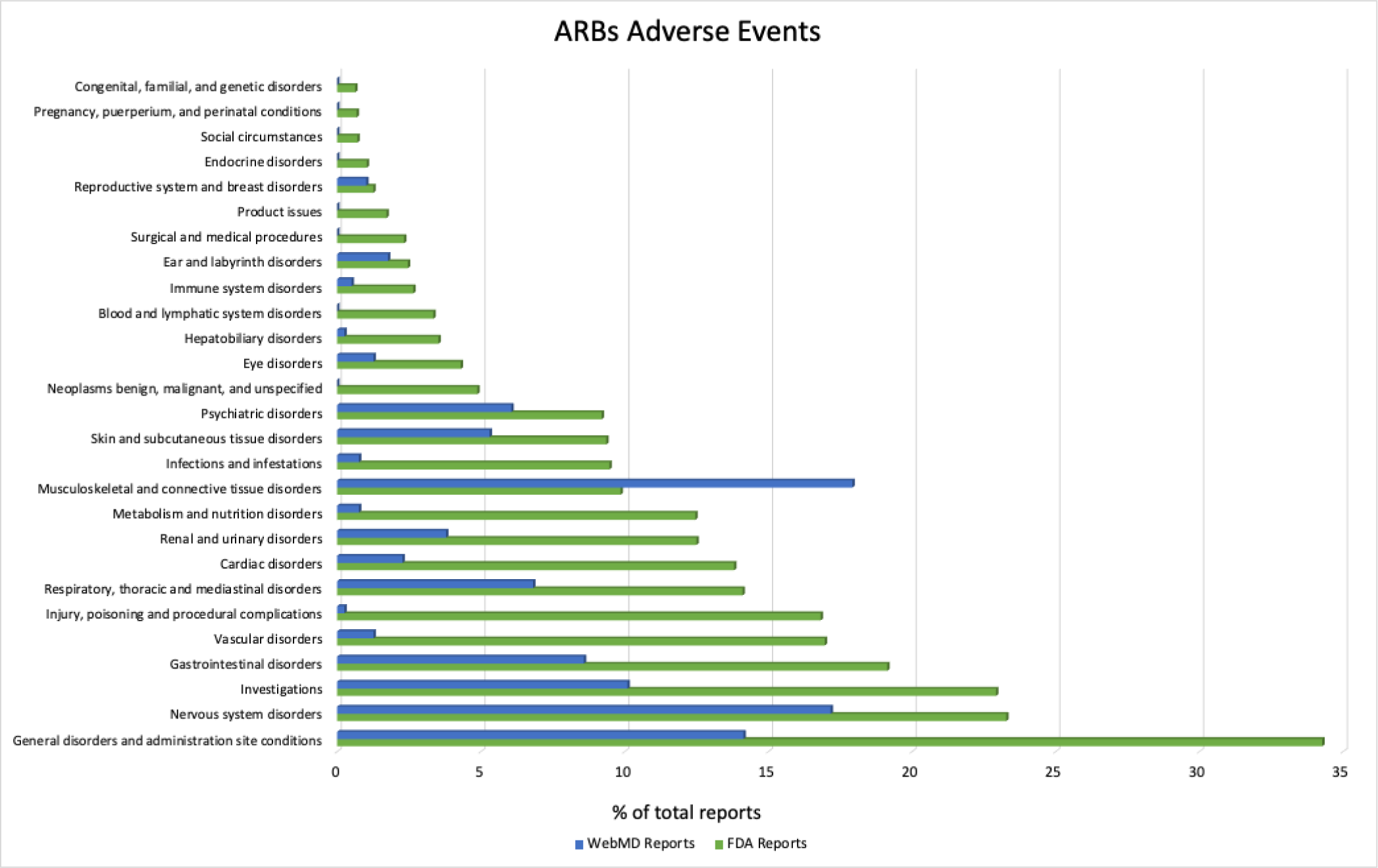
ARBs adverse event SOC category comparison of FDA (n=759,742) and WebMD (n=396) reports

**Figure 3.**
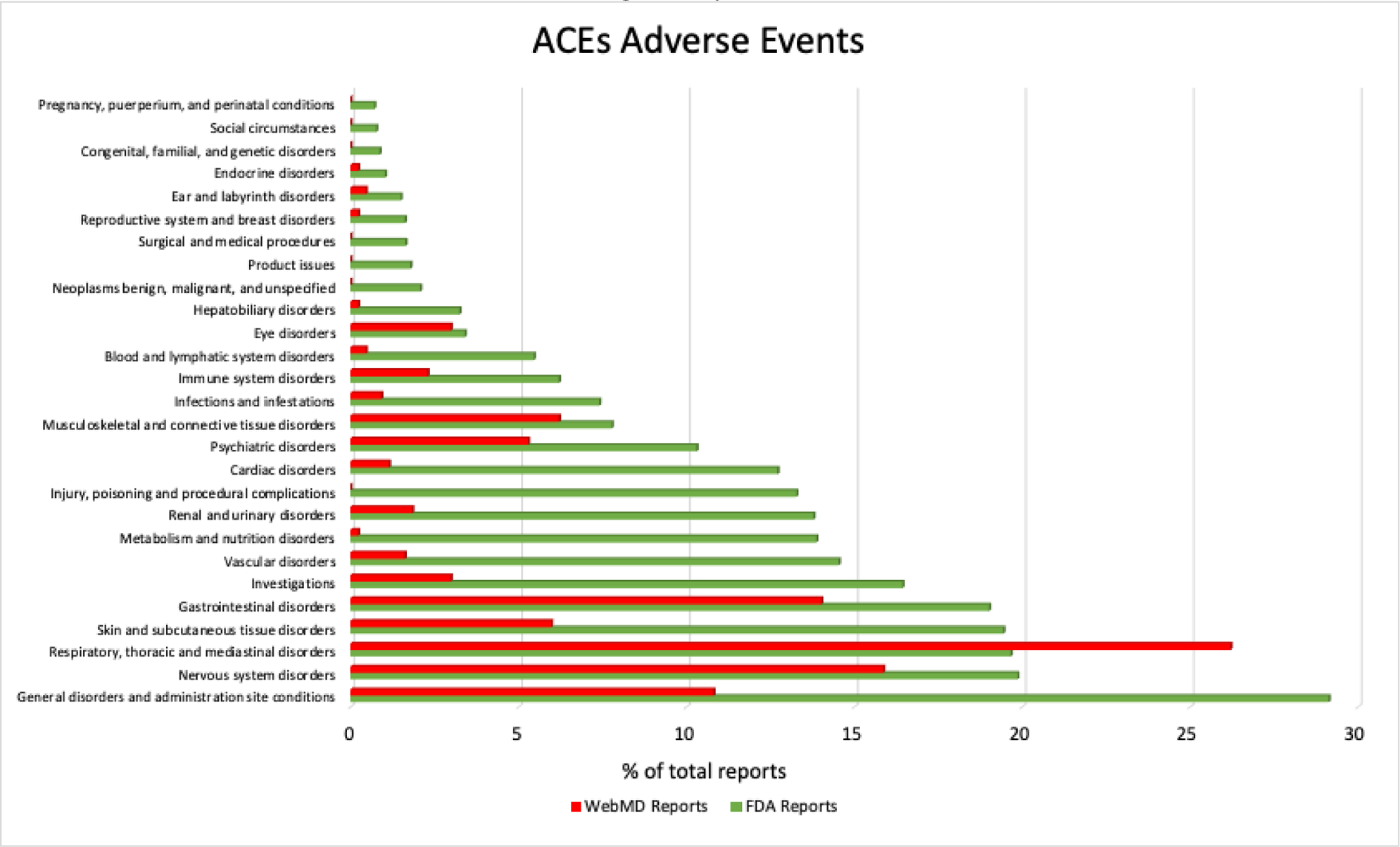
ACEIs adverse event SOC category comparison of FDA (n=512,196) and WebMD (n=435) reports

Percentage point difference (as a fraction of the total) was used to further compare FDA and WebMD data for both ARBs and ACEIs datasets. For the ARBs, most differences were not statistically significant with the exception of ‘Vascular disorders’ (−5.4 % points difference, p=0.010) and ‘Injury, poisoning and procedural complications’ (−6.4 % points difference, p=0.002) being reported as a lower proportion of the total in WebMD data than FDA data and ‘Nervous system disorders’ (8 % points difference, p=0.001) and ‘Musculoskeletal and connective tissue disorders (14 % points difference, p=0.000) being reported as a higher proportion of the total in WebMD data than FDA data. For the ACEIs, the only statistically significant differences were ‘Metabolism and nutrition disorders’ (−5.4 % points difference, p=0.001) and ‘Injury, poisoning and procedural complications’ (−5.4 % points difference, p=0.001) being reported as a lower proportion of the total in WebMD data than FDA data and ‘Nervous system disorders’ (7.8 % points difference, p=0.000), ‘Respiratory, thoracic and mediastinal disorders’ (18.3 % points difference, p=0.000) and ‘Gastrointestinal disorders’ (6.3 % points difference, p=0.001) being reported as a higher proportion of the total in WebMD data than FDA data.

**Table 1.** ARBs adverse event rank comparison across WebMD, FDA, and MHRA data.

| MedDRA Primary SOC Code | Number of WebMD patients taking ARBs reporting adverse event(s) from the primary SOC code | Rank order of **WebMD** patients taking ARBs reporting adverse event(s) from the primary SOC code | Rank order of **FDA** report data | Rank order of **MHRA** report data |
| --- | --- | --- | --- | --- |
| Musculoskeletal and connective tissue disorders | 71 | 1 | 11 | 7 |
| Nervous system disorders | 68 | 2 | 2 | 2 |
| General disorders and administration site conditions | 56 | 3 | 1 | 1 |
| Investigations | 40 | 4 | 3 | 6 |
| Gastrointestinal disorders | 34 | 5 | 4 | 3 |
| Respiratory, thoracic, and mediastinal disorders | 27 | 6 | 7 | 5 |
| Psychiatric disorders | 24 | 7 | 14 | 10 |
| Skin and subcutaneous tissue disorders | 21 | 8 | 13 | 4 |
| Renal and urinary disorders | 15 | 9 | 9 | 8 |
| Cardiac disorders | 9 | 10 | 8 | 12 |

*Red= More than two ranks higher in WebMD than federal data, Green= WebMD and federal data within 2 ranks, Yellow= More than two ranks higher in federal data than WebMD*

**Table 2.** ACEIs adverse event rank comparison across WebMD, FDA, and MHRA data.

| MedDRA Primary SOC Code | Number of WebMD patients taking ACEIs reporting adverse event(s) from the primary SOC code | Rank order of WebMD patients taking ACEIs reporting adverse event(s) from the primary SOC code | Rank order of FDA report data | Rank order of MHRA report data |
| --- | --- | --- | --- | --- |
| Respiratory, thoracic, and mediastinal disorders | 114 | 1 | 3 | 1 |
| Nervous system disorders | 69 | 2 | 2 | 5 |
| Gastrointestinal disorders | 61 | 3 | 5 | 3 |
| General disorders and administration site conditions | 47 | 4 | 1 | 4 |
| Musculoskeletal and connective tissue disorders | 27 | 5 | 13 | 9 |
| Skin and subcutaneous tissue disorders | 26 | 6 | 4 | 2 |
| Psychiatric disorders | 23 | 7 | 12 | 7 |
| Investigations | 13 | 8 | 6 | 10 |
| Eye disorders | 13 | 8 | 17 | 14 |
| Immune system disorders | 10 | 10 | 15 | 16 |

*Red= More than two ranks higher in WebMD than federal data, Green= WebMD and federal data within 2 ranks, Yellow= More than two ranks higher in federal data than WebMD*

### 3.3 Non-persistence and General Quote Analysis

Posts were identified as confirmed non-persistence (discontinuation outside of medical advice) cases when they were coded as a patient-directed change and as a “no” under the healthcare consultation column. Possible non-persistence cases are coded as “unsure” in the healthcare consultation column. There were 18/142 (13%) confirmed and 18-77/142 (13%-54%) possible cases of non-persistence in the ARBs dataset and 26/187 (14%) confirmed and 26-91/187 (14%-49%) possible cases of non-persistence in the ACEIs dataset.

Most of the patients posting on WebMD about discontinuation or switching due to adverse events did so for one or a combination of the following three reasons: (1) to express concern or anger about an adverse event they experienced, (2) to warn others about the “dangers” of a drug that they have taken, and/or (3) to express a realization that they “are not alone” in the experience of adverse events they have had. Many posts in the confirmed and possible non-persistence categories expressed a lack of patient-physician trust, stating comments like: “doctors don’t care,” “all [physicians] want to do is shove a pill down your throat,” or “my doctor does not believe any med causes side effects.” Others stated that their physician “showed almost no concern” or “never warned [them]” about certain side effects.

## 4 Discussion

The major reason for the discontinuation of ARBs and ACEIs, with or without healthcare consultation, expressed through WebMD was the experience of adverse events. For ARBs, ‘Vascular disorders’ and ‘Injury, poisoning and procedural complications’ SOC categories were reported less in WebMD data than FDA data, and ‘Nervous system disorders’ and ‘Musculoskeletal and connective tissue disorders’ were reported more in WebMD data than FDA data. For ACEIs, ‘Metabolism and nutrition disorders’ and ‘Injury, poisoning and procedural complications’ were reported less in WebMD data than FDA data, and ‘Nervous system disorders’, ‘Respiratory, thoracic and mediastinal disorders’, and ‘Gastrointestinal disorders’ were reported more in WebMD data than FDA data.

Previous studies about using FAERS data have found that it is a beneficial tool for pharmacovigilance; however, it is not devoid of reporting bias, and its data must be interpreted with the consideration of various confounding variables [27, 28]. Firstly, there is no way to control the date of the data being extracted as the online database only performs a search of all FAERS data since 1968, making unmatching timelines a confounding variable when using FAERS as a comparison. Date-specific data could be extracted using quarterly reports, but time constraints did not make this feasible for this study. Secondly, there has been a continual two-fold increase in the amount of serious adverse events being reported to the FDA database from 2006 to 2014 [27], which supports the increased number of reports to the FDA in ‘Vascular disorders’ for ARBs, ‘Metabolism and nutrition disorders’ for ACEIs, and ‘Injury, poisoning and procedural complications for both ARBs and ACEIs. These SOC categories either require medical tests and diagnoses or are related to medication misuse that are more conducive to formal FDA reporting than to WebMD reporting. Reporting trends tend to vary with time and are influenced by a variety of external factors such as negative news media coverage, litigation, and views regarding the government and the FDA [27–29]. A 2017 study found that only approximately 62.5% of U.S. adults trust the FDA [30], and trust in the FDA and CDC is currently at an all-time low due to the COVID-19 pandemic [31]. The decrease in trust in the FDA could lower reports of adverse events through the FAERS, making online drug review sites such as WebMD a source of information that could fill a significant gap, validate FAERS data, or even provide insight into the confounding variables affecting adverse events reports. In 2015, Golder et al. found that mentions of mild or symptomatic adverse events were more prevalent in social media data than in other data sources [33]. WebMD is a more informal platform that may motivate patients to report and converse about less drastic or more chronic adverse effects [33] like muscle/joint pains (Musculoskeletal and connective tissue disorders’) in the case of ARBs, mild coughs (‘Respiratory, thoracic and mediastinal disorders’) or abdominal discomfort (‘Gastrointestinal disorders’) in the case of ACEIs, or dizziness (‘Nervous system disorders’) in the case of both ARBs and ACEIs.

An important note to take into account is that the rank of the Musculoskeletal and connective tissue disorders SOC category was much higher in the WebMD reports than in both federal databases analyzed. There is little research about reporting rates for Musculoskeletal and connective tissue adverse events; however, a prior study about the ability of a website for patient support (Askapatient.com) to detect adverse events for atorvastatin and sibutramine showed that musculoskeletal adverse events were reported approximately 40% more on social media than the FAERS [34]. Moreover, a study focusing on how media attention affects the reporting of adverse events found no changes in reporting except for a significant increase in reports of Musculoskeletal and connective tissue disorders SOC after a media broadcast about statins and adverse reactions [35]. This outlying increase in reporting may indicate that musculoskeletal adverse reaction reporting on social media may be more susceptible to changes in media coverage or other confounding variables. Furthermore, FDA data can only provide an indication of overall adverse events experienced, while this study aims to identify the adverse events that lead to a change in medication.

The comments of confirmed nonadherence posts frequently explained that the experience of adverse events and a lack of trust in the relationship with their physician led to the decision of discontinuing their medication. The comments mentioned that their physicians either had not warned the patient of the adverse events they experienced, or that the physician did not think the adverse events experienced by the patient were related to the antihypertensive medication. Insight into patient perspectives on ACEIs and ARBs adverse events may allow physicians and other healthcare personnel to better understand and address these concerns. This knowledge will also discourage nonadherence by helping build a stronger patient-physician relationship that will encourage patients to seek healthcare consultations whenever they have concerns.

This study shows the promise of WebMD mining in the study of hypertensive medication nonadherence and adverse events. The results provide insight into patient concerns about hypertensive medication treatment plans, and understanding these concerns may be just as important as studying the pharmacological adverse effects of antihypertensive medications. Many of the adverse events reported for antihypertensive medications are ambiguous (headaches, fatigue, dizziness, etc.) as to whether they are due to the medication or not. Either way, they can drive medication discontinuation and nonadherence.

### 4.1 Limitations and Future Areas of Study

The results of the study are limited by the small sample size of WebMD posts that were analyzed. However, after reviewing 300 ARBs posts and 300 ACEIs posts, some data-saturation was reached. Moreover, because the initial corpus of ARBs posts was more extensive than the ACEIs corpus, the first 300 ARBs posts are more recent than the first 300 ACEIs posts. Although time did not seem to have a significant impact on the annotation data collected, a larger sample size would be needed in further research. Both FDA or MHRA reports and WebMD posts are of a voluntary nature and not representative of all patients taking antihypertensive drugs; therefore, reports may be skewed towards those with the interest, time, and means to report or disclose the reasons for their medication discontinuation. Previous studies have found that women are more likely than men to report adverse drug events for both ACEIs and ARBs [32], and the current study reflects that. Additionally, social media posting may slightly skew results towards middle-age participants; however, more in-depth analyses of WebMD reporting demographics must be studied to determine this.

### 4.2 Implications in Health Care

The findings of this study demonstrate the potential of the analysis of WebMD posts to understand patient perspectives on their medications and their patient-physician relationships. It is important to not only understand if medications are physiologically causing patients adverse effects, but also understand the perspectives and concerns that are causing patients to discontinue their medication without a healthcare consultation. When used in combination with FDA data, WebMD has the potential to provide a clearer picture of adverse events. It can be especially useful for chronic medications or treatments that must be continually studied since adverse events may arise only after a long time of taking medication. Similarities with FDA data could help increase the validity of information from either source, and differences could help increase the breadth of adverse events that could be studied and analyzed.

## 5 Conclusion

This study builds upon the Golder et al. 2022 study of statin discontinuation through WebMD. Hypertension is the leading cause of heart disease in the world [12–15]; therefore, obtaining a thorough understanding of antihypertensive medication discontinuation and adverse events (especially when nonadherence is involved) could be significantly beneficial.

The study provides valuable foundational research that addresses a population with significant rates of nonadherence. Comparisons to other hypertensive medications such as calcium channel blockers, beta blockers, and diuretics could reveal other facets of chronic antihypertensive medication treatment nonadherence. Further research is needed to verify the reliability of the WebMD datasource with larger sample sizes, more classes of antihypertensive medication, and more comparisons to other pharmacovigilance methods.

Online drug review platforms, such as WebMD, have the potential to provide both quantitative data about demographics and reasons for medication discontinuation and qualitative data about patient perspectives and opinions of adverse events, insurance, and other problems with certain prescriptions.

## Data Availability

All data produced in the present study are available upon reasonable request to the authors

## Funding

This work was supported by National Institutes of Health (NIH) National Library of Medicine under grant number NIH NLM 1R01. NIH National Library of Medicine funded this research but were not involved in the design and conduct of the study; collection, management, analysis, and interpretation of the data; preparation, review, or approval of the manuscript; and decision to submit the manuscript for publication. CM, SG, and KO had full access to all the data in the study and take responsibility for the integrity of the data and the accuracy of the data analysis.

## Declarations

### Conflicts of Interests

The Authors declare no Competing Financial or Non-Financial Interests.

### Ethics Approval

All data used in this study were collected according to the WebMD terms of use and were publicly available at the time of collection and analysis. We have an institutional review board certificate of exemption from the University of Pennsylvania.

### Consent to Participate

Not applicable.

### Consent for Publication

Not applicable.

### Availability of Data and Material

The drug reviews included in this project are publicly available on WebMD. The coding underpinning this article will be shared on reasonable request to the corresponding author.

### Author Contributions

The authors confirm contribution to the paper as follows: study conception and design: CM, SG, DW, GG.; data collection: DW; analysis and interpretation of results: CM, SG, KO. Draft manuscript preparation: CM. All authors reviewed the results and approved the final version of the manuscript. Acquisition of funding: GG.

## Supplementary Materials

**Table 1.** List of medications included in WebMD search.

|  | Generic | Brand |
| --- | --- | --- |
| Angiotensin II Receptor Blockers (ARBs) | azilsartan medoxomil | Edarbi |
|  | candesartan | Atacand |
|  | eprosartan | Teveten |
|  | irbesartan | Avapro |
|  | losartan | Cozaar |
|  | olmesartan | Benicar |
|  | telmisartan | Micardis |
|  | valsartan | Diovan<br>Prexxartan |
| Angiotensin-Converting Enzyme Inhibitors (ACEIs) | benazepril | Lotensin |
|  | captopril | Capoten |
|  | enalapril | Enalaprilat<br>Vasotec<br>Epaned |
|  | fosinopril | Monopril |
|  | lisinopril | Zestril<br>Prinivil |
|  | moexipril | Univasc |
|  | perindopril | Aceon |
|  | quinapril | Accupril |
|  | ramipril | Altace |
|  | trandolapril | Mavik |

**Table 2.**
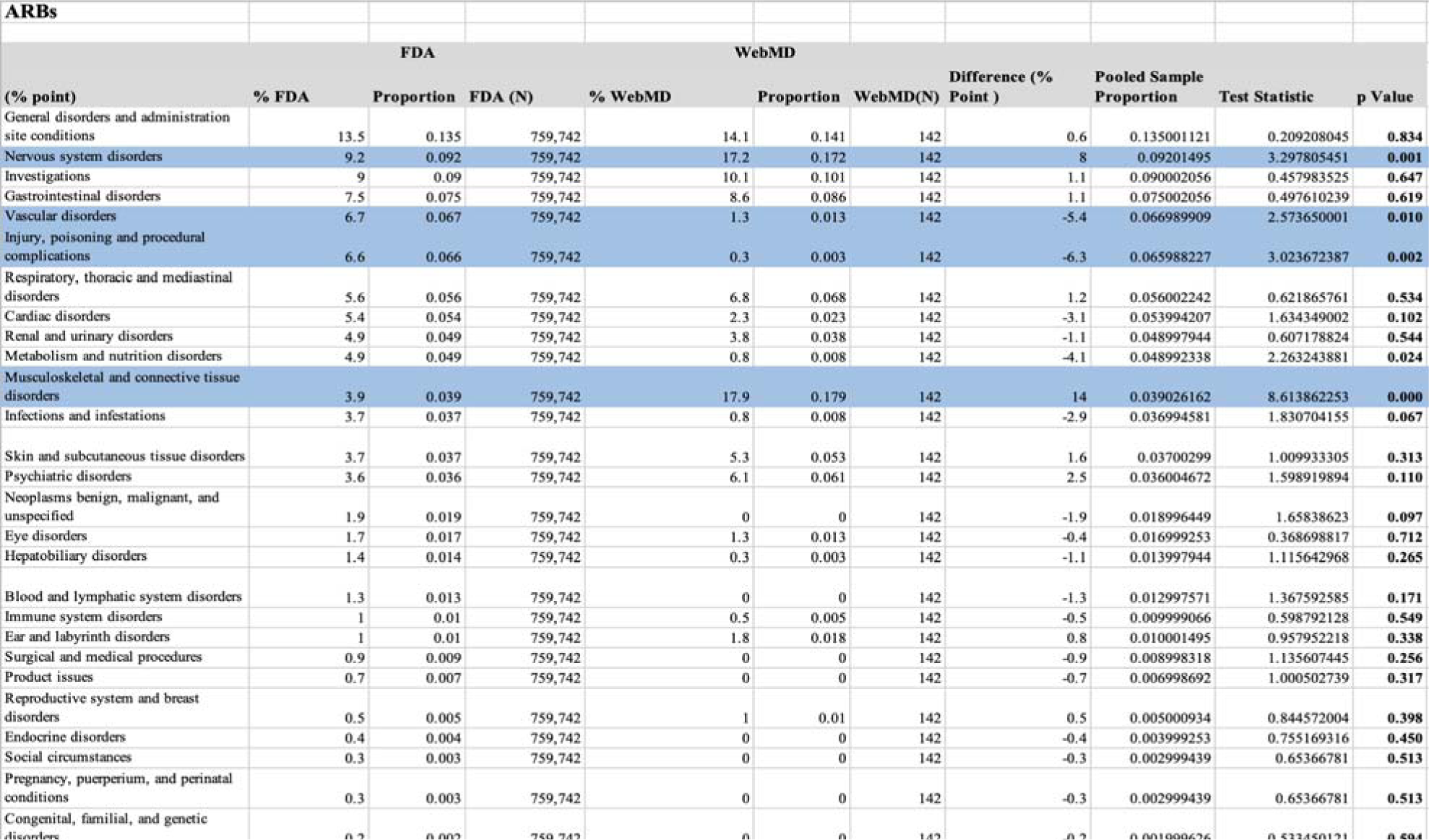
ARBs p-values and data analysis for WebMD and FDA SOC category comparison.

**Table 3.** ACEIs p-values and data analysis for WebMD and FDA SOC category comparison.

| ACEIs |  |  |  |  |  |  |  |  |  |  |
| --- | --- | --- | --- | --- | --- | --- | --- | --- | --- | --- |
|  | FDA |  |  | WebMD |  |  |  |  |  |  |
| (% point) | % FDA | Proportion | FDA (N) | % WebMD | Proportion | WebMD(N) | Difference (% Point ) | Pooled Sample Proportion | Test Statistic | p Value |
| General disorders and administration site conditions | 11.77 | 0.1177 | 512196 | 10.8 | 0.108 | 187 | -1.0 | 0.11769646 | 0.411549958 | **0.681** |
| Nervous system disorders | 8.02 | 0.0802 | 512196 | 15.86 | 0.1586 | 187 | 7.8 | 0.080228613 | 3.945963026 | **0.000** |
| Respiratory, thoracic and mediastinal disorders | 7.94 | 0.0794 | 512196 | 26.21 | 0.2621 | 187 | 18.3 | 0.079466678 | 9.235657067 | **0.000** |
| Skin and subcutaneous tissue disorders | 7.85 | 0.0785 | 512196 | 5.98 | 0.0598 | 187 | -1.9 | 0.078493175 | 0.950644015 | **0.342** |
| Gastrointestinal disorders | 7.68 | 0.0768 | 512196 | 14.02 | 0.1402 | 187 | 6.3 | 0.076823139 | 3.254934983 | **0.001** |
| Investigations | 6.64 | 0.0664 | 512196 | 2.99 | 0.0299 | 187 | -3.7 | 0.066386679 | 2.004520082 | **0.045** |
| Vascular disorders | 5.87 | 0.0587 | 512196 | 1.61 | 0.0161 | 187 | -4.3 | 0.058684453 | 2.478118059 | **0.013** |
| Metabolism and nutrition disorders | 5.6 | 0.056 | 512196 | 0.23 | 0.0023 | 187 | -5.4 | 0.055980402 | 3.19379704 | **0.001** |
| Renal and urinary disorders | 5.57 | 0.0557 | 512196 | 1.84 | 0.0184 | 187 | -3.7 | 0.055686387 | 2.223912647 | **0.026** |
| Injury, poisoning and procedural complications | 5.37 | 0.0537 | 512196 | 0 | 0 | 187 | -5.4 | 0.053680402 | 3.257534537 | **0.001** |
| Cardiac disorders | 5.14 | 0.0514 | 512196 | 1.15 | 0.0115 | 187 | -4.0 | 0.051385438 | 2.470867796 | **0.013** |
| Psychiatric disorders | 4.16 | 0.0416 | 512196 | 5.29 | 0.0529 | 187 | 1.1 | 0.041604124 | 0.773711982 | **0.439** |
| Musculoskeletal and connective tissue disorders | 3.14 | 0.0314 | 512196 | 6.21 | 0.0621 | 187 | 3.1 | 0.031411204 | 2.406400447 | **0.016** |
| Infections and infestations | 2.99 | 0.0299 | 512196 | 0.92 | 0.0092 | 187 | -2.1 | 0.029892445 | 1.661962495 | **0.097** |
| Immune system disorders | 2.51 | 0.0251 | 512196 | 2.3 | 0.023 | 187 | -0.2 | 0.025099234 | 0.183548313 | **0.854** |
| Blood and lymphatic system disorders | 2.21 | 0.0221 | 512196 | 0.46 | 0.0046 | 187 | -1.8 | 0.022093613 | 1.627786989 | **0.104** |
| Eye disorders | 1.37 | 0.0137 | 512196 | 2.99 | 0.0299 | 187 | 1.6 | 0.013705912 | 1.905019224 | **0.057** |
| Hepatobiliary disorders | 1.31 | 0.0131 | 512196 | 0.23 | 0.0023 | 187 | -1.1 | 0.013096058 | 1.298845668 | **0.194** |
| Neoplasms benign, malignant, and unspecified | 0.84 | 0.0084 | 512196 | 0 | 0 | 187 | -0.8 | 0.008396934 | 1.258610979 | **0.208** |
| Product issues | 0.72 | 0.0072 | 512196 | 0 | 0 | 187 | -0.7 | 0.007197372 | 1.16454317 | **0.244** |
| Surgical and medical procedures | 0.66 | 0.0066 | 512196 | 0 | 0 | 187 | -0.7 | 0.006597591 | 1.11462856 | **0.265** |
| Reproductive system and breast disorders | 0.65 | 0.0065 | 512196 | 0.23 | 0.0023 | 187 | -0.4 | 0.006498467 | 0.714662664 | **0.475** |
| Ear and labyrinth disorders | 0.6 | 0.006 | 512196 | 0.46 | 0.0046 | 187 | -0.1 | 0.005999489 | 0.247867244 | **0.804** |
| Endocrine disorders | 0.42 | 0.0042 | 512196 | 0.23 | 0.0023 | 187 | -0.2 | 0.004199307 | 0.401716931 | **0.688** |
| Congenital, familial, and genetic disorders | 0.35 | 0.0035 | 512196 | 0 | 0 | 187 | -0.4 | 0.003498723 | 0.810430721 | **0.418** |
| Social circumstances | 0.31 | 0.0031 | 512196 | 0 | 0 | 187 | -0.3 | 0.003098869 | 0.762562783 | **0.446** |
| Pregnancy, puerperium, and perinatal conditions | 0.29 | 0.0029 | 512196 | 0 | 0 | 187 | -0.3 | 0.002898942 | 0.737479943 | **0.461** |

## Annotation Guidelines

### Project Background

Medication nonadherence refers to when a patient does not adhere to their prescription regimen. Nonadherence to anti-hypertensive medication can have severe consequences, and it has become a serious issue as approximately 60% of controlled blood pressure patients are nonadherent to anti-hypertensive medication. Researching and monitoring the reasons behind anti-hypertensive medication non-adherence is essential to finding an effective solution for it. In this project, we will be manually annotating a set of WebMD comments for ACE inhibitors and Angiotensin II Receptor Blockers that indicate a medication change.

### General Annotation Instructions

1. <u>Medication change column:</u> if the comment does not mention a medication change, it should be coded with a ‘0’; if is mentions a medication change, it should be coded with a ‘d’ for discontinuation, ‘s’ for medication switch, or ‘c’ for a dosage change
2. <u>Health care consultation column:</u> coded with ‘yes’, ‘no’, or ‘unsure’ depending on whether the author appears to have had healthcare direction to change medication or prescription regimen
3. <u>Patient/healthcare directed column:</u> comment should be coded with a ‘p’ if the change appeared to be mostly directed by the author/patient or with an ‘h’ if the change was prompted by a healthcare professional
4. <u>Reason for change columns</u>: one or two reasons from the dropdown list of reasons for the change (1: adverse effects, 2: cost/insurance, 3: not working, 4: not necessary, 5: other) should be selected for each comment that is a positive for medication change
5. <u>Adverse effects extraction:</u> if one of the reasons for nonadherence in the previous column was ‘adverse effects’, then the annotator should extract the mentions of specific adverse effects from the comment and insert them into this column
6. <u>Days to adverse effects:</u> if mentioned, amount of time (in days) after start of medication regimen at which adverse effects started should be recorded in this column
7. <u>Rechallenge and “Side Effects back?” columns:</u> if comment mentions retrying a certain medication or medication regimen, the rechallenge column should be coded with a ‘yes’; “side effects back?” column should be coded with a ‘yes’ or a ‘no’ depending on whether adverse effects return after restarting certain medication
8. <u>Notes column</u>: any additional interesting or recurring details mentioned in the comment should be noted in this column

### What should be coded as a medication change?

1. Any mention of a change in medication regimen (a discontinue, increase/decrease, or exchange of medication)
2. Mentioned medication change should relate to a personal experience of the author

### What should be coded as an adverse effect?

1. Effects originate from taking the specific drug the comment was made for, and they were not due to taking too much of the drug
2. Author personally experiences the effects
3. Effect can be coded to a medical term in the dropdown menu even if it is in colloquial terms

EXAMPLE 1: *Accupril*

“I had been on Accupril or quinapril 10 for at least 25 years. I had no problems and it kept the blood pressure under control. Last month I was rushed to the hosp. with swelling of my tongue which I now know is a side effect. It can happen any time. It happened again the other night. Drs. advised it can stay in system for 6 weeks. I have changed BP medicine.”

| Medication Change | Health care consultation | Patient/healthcare directed | Reason for change | Adverse effects extraction | Days to adverse effect | Rechallenge | Side effects back? |
| --- | --- | --- | --- | --- | --- | --- | --- |
| s | yes | p | adverse effects | Swelling of tongue | 9125** | no |  |

\*\*25 years written as days

EXAMPLE 2: Losartan Potassium

“After reading other comments, I see I am not alone in the profuse sweating I experience every time I do anything physical. Also experiencing more headaches than normal and noticing hands seem swollen in the mornings. Also have some strange muscle aches and pains that do not seem to come from my exercise routine. My doctor does not believe any med causes side effects so I am sure he will not believe he needs to change this medicine for me. Will just quit taking it and hope for the best”

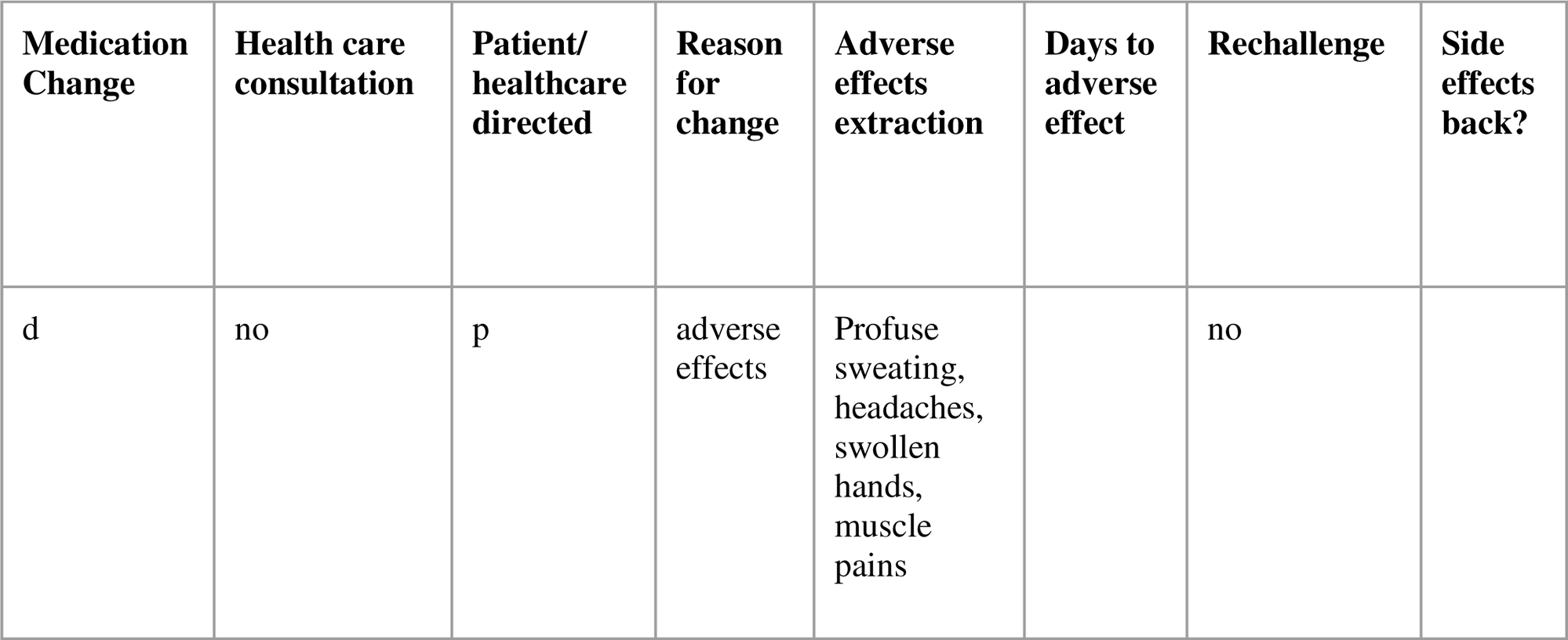

## Notes

### Competing Interest Statement

The authors have declared no competing interest.

## References

(1) Hugtenburg, J. G., Timmers, L., Elders, P. J., Vervloet, M., & van Dijk, L. (2013). Definitions, variants, and causes of nonadherence with medication: a challenge for tailored interventions. Patient preference and adherence, 7, 675.

(2) Onishi, T., Weissenbacher, D., Klein, A., O’Connor, K., & Gonzalez, G. (2018, October). Dealing with medication non-adherence expressions in twitter. In Proceedings of the 2018 EMNLP Workshop SMM4H: The 3rd Social Media Mining for Health Applications Workshop & Shared Task (pp. 32–33).

(3) Walsh, C. A., Cahir, C., Tecklenborg, S., Byrne, C., Culbertson, M. A., & Bennett, K. E. (2019). The association between medication non-adherence and adverse health outcomes in ageing populations: A systematic review and meta-analysis. British journal of clinical pharmacology, 85(11), 2464–2478.

(4) Cramer, J. A., Benedict, A., Muszbek, N., Keskinaslan, A., & Khan, Z. M. (2008). The significance of compliance and persistence in the treatment of diabetes, hypertension and dyslipidaemia: a review. International journal of clinical practice, 62(1), 76–87.

(5) Zhou, J., Han, J., Nutescu, E. A., Patel, P. R., Sweiss, K., & Calip, G. S. (2019). Discontinuation and Nonadherence to Medications for Chronic Conditions after Hematopoietic Cell Transplantation: A 6lJYear Propensity Score–Matched Cohort Study. Pharmacotherapy: The Journal of Human Pharmacology and Drug Therapy, 39(1), 55–66.

(6) Jüngst, C., Gräber, S., Simons, S., Wedemeyer, H., & Lammert, F. (2019). Medication adherence among patients with chronic diseases: a survey-based study in pharmacies. QJM: An International Journal of Medicine, 112(7), 505–512.

(7) Greene, M., Yan, T., Chang, E., Hartry, A., Touya, M., & Broder, M. S. (2018). Medication adherence and discontinuation of long-acting injectable versus oral antipsychotics in patients with schizophrenia or bipolar disorder. Journal of Medical Economics, 21(2), 127–134.

(8) Beeftink, M. M., Van Der Sande, N. G., Bots, M. L., Doevendans, P. A., Blankestijn, P. J., Visseren, F. L., … & Spiering, W. (2017). Safety of temporary discontinuation of antihypertensive medication in patients with difficult-to-control hypertension. Hypertension, 69(5), 927–932.

(9) van der Wardt, V., Harrison, J. K., Welsh, T., Conroy, S., & Gladman, J. (2017). Withdrawal of antihypertensive medication: a systematic review. Journal of hypertension, 35(9), 1742.

(10) Jimmy, B., & Jose, J. (2011). Patient medication adherence: measures in daily practice. Oman medical journal, 26(3), 155.

(11) Thio, S. L., Nam, J., van Driel, M. L., Dirven, T., & Blom, J. W. (2018). Effects of discontinuation of chronic medication in primary care: a systematic review of deprescribing trials. British Journal of General Practice, 68(675), e663–e672.

(12) Abegaz, T. M., Shehab, A., Gebreyohannes, E. A., Bhagavathula, A. S., & Elnour, A. A. (2017). Nonadherence to antihypertensive drugs: a systematic review and meta-analysis. Medicine, 96(4).

(13) Centers for Disease Control and Prevention (CDC). Hypertension Cascade: Hypertension Prevalence, Treatment and Control Estimates Among US Adults Aged 18 Years and Older Applying the Criteria From the American College of Cardiology and American Heart Association’s 2017 Hypertension Guideline—NHANES 2013–2016. Atlanta, GA: US Department of Health and Human Services; 2019.

(14) Gallagher, B. D., Muntner, P., Moise, N., Lin, J. J., & Kronish, I. M. (2015). Are two commonly used self-report questionnaires useful for identifying antihypertensive medication non-adherence?. Journal of hypertension, 33(5), 1108.

(15) file:///Users/cristinamicale/Downloads/HYPERTENSIONAHA.116.07720.pdf

(16) Cooke, C. E., Xing, S., Gale, S. E., & Peters, S. (2021). Initial non-adherence to antihypertensive medications in the United States: a systematic literature review. Journal of Human Hypertension, 1–11.

(17) Chang, T. E., Ritchey, M. D., Park, S., Chang, A., Odom, E. C., Durthaler, J., … & Loustalot, F. (2019). National rates of nonadherence to antihypertensive medications among insured adults with hypertension, 2015. Hypertension, 74(6), 1324–1332.

(18) Charalabopoulos, K., Charalabopoulos, A., Papalimneou, V., Kiortsis, D., Dimicco, P., Kostoula, O. K., & Karachalios, G. N. (2005). Consequences of the discontinuation of antihypertensive treatment in successfully treated patients. International journal of clinical practice, 59(6), 704–708.

(19) Weissenbacher, D., Ge, S., Klein, A., O’Connor, K., Gross, R., Hennessy, S., & Gonzalez-Hernandez, G. (2021). Active neural networks to detect mentions of changes to medication treatment in social media. Journal of the American Medical Informatics Association, 28(12), 2551–2561.

(20) Struthers, A. D., Anderson, G., MacFadyen, R. J., Fraser, C., & MacDonald, T. M. (2001). Nonadherence with ACE inhibitors is common and can be detected in clinical practice by routine serum ACE activity. Congestive Heart Failure, 7(1), 43–50.

(21) Herman, L. L., Padala, S. A., Ahmed, I., & Bashir, K. (2017). Angiotensin converting enzyme inhibitors (ACEI).

(22) Hill, R. D., & Vaidya, P. N. (2019). Angiotensin II Receptor Blockers (ARB).

(23) Xie, J., Zeng, D. D., & Marcum, Z. A. (2017). Using deep learning to improve medication safety: the untapped potential of social media. Therapeutic Advances in Drug Safety, 8(12), 375–377.

(24) Lam, W. Y., & Fresco, P. (2015). Medication adherence measures: an overview. BioMed research international, 2015.

(25) Golder, S., Weissenbacher, D., O’Connor, K., Hennessy, S., Gross, R., & Hernandez, G. G. (2022). Patient-Reported Reasons for Switching or Discontinuing Statin Therapy: A Mixed Methods Study Using Social Media. Drug Safety, 45(9), 971–981.

(26) Weissenbacher, D., et al. Active neural networks to detect mentions of changes to medication treatment in social media. J Am Med Inform Assoc 28, 2551–2561 (2021).

(27) Sonawane, K. B., Cheng, N., & Hansen, R. A. (2018). Serious adverse drug events reported to the FDA: analysis of the FDA Adverse Event Reporting System 2006-2014 database. Journal of managed care & specialty pharmacy, 24(7), 682–690.

(28) Monnot, A. D., Fung, E. S., Compoginis, G. S., & Towle, K. M. (2021). An evaluation of the FDA adverse event reporting system and the potential for reporting bias. Journal of Cosmetic Dermatology, 20(6), 1849–1854.

(29) Moore TJ, Cohen MR, Furberg CD. Serious adverse drug events reported to the Food and Drug Administration, 1998-2005. Arch Intern Med. 2007; 167(16):1752–59.

(30) Kowitt, S. D., Schmidt, A. M., Hannan, A., & Goldstein, A. O. (2017). Awareness and trust of the FDA and CDC: Results from a national sample of US adults and adolescents. PloS one, 12(5), e0177546.

(31) Nahum, A., Drekonja, D. M., & Alpern, J. D. (2021, February). The erosion of public trust and SARS-CoV-2 vaccines—more action is needed. In Open forum infectious diseases (Vol. 8, No. 2, p. ofaa657). US: Oxford University Press.

(32) Rydberg, D. M., Mejyr, S., Loikas, D., Schenck-Gustafsson, K., von Euler, M., & Malmström, R. E. (2018). Sex differences in spontaneous reports on adverse drug events for common antihypertensive drugs. European journal of clinical pharmacology, 74(9), 1165–1173.

(33) Golder, S., Norman, G., & Loke, Y. K. (2015). Systematic review on the prevalence, frequency and comparative value of adverse events data in social media. British journal of clinical pharmacology, 80(4), 878–888.

(34) Duh, M. S., Cremieux, P., Audenrode, M. V., Vekeman, F., Karner, P., Zhang, H., & Greenberg, P. (2016). Can social media data lead to earlier detection of druglJrelated adverse events?. Pharmacoepidemiology and drug safety, 25(12), 1425–1433.

(35) van Hunsel, F., van Puijenbroek, E., de JonglJvan den Berg, L., & van Grootheest, K. (2010). Media attention and the influence on the reporting odds ratio in disproportionality analysis: an example of patient reporting of statins. Pharmacoepidemiology and drug safety, 19(1), 26–32.

